# Maternal HIV retesting during pregnancy and postpartum among high-risk populations in Kenya, South Africa, and Ukraine: Cost-effectiveness of preventing vertical transmission

**DOI:** 10.1101/2025.04.08.25325413

**Authors:** David M. Coomes, Julianne Meisner, D Allen Roberts, Patricia Rodriguez, Morkor Newman Owiredu, Monisha Sharma, Alexey Bobrik, Arkadii Vodianyk, Amanda Williams, George Githuka, Thato Chidarikire, Ruanne V. Barnabas, Shiza Farid, Shaffiq Essajee, Muhammad S. Jamil, Rachel Baggaley, Cheryl C. Johnson, Alison L. Drake

**Author notes:** Corresponding author: Alison L. Drake University of Washington, 3980 15th Ave NE Rm 884, Box 351620, Seattle, WA 98195, USA.

## Abstract

**Introduction:** Key populations, including female sex workers (FSW), people who inject drugs (PWID), and people in serodiscordant partnerships, experience higher HIV incidence compared to the general population. Maternal HIV retesting, particularly during late pregnancy, helps detect new infections and prevent vertical HIV transmission, but optimal testing schedules among key populations are unknown.

**Methods:** We used a Markov model to estimate the health and economic impacts of maternal HIV retesting on vertical HIV transmission outcomes among FSW and PWID in Kenya, South Africa, and pre-war Ukraine as well as among pregnant people in serodiscordant partnerships in Kenya and South Africa. We calculated incremental cost-effectiveness ratios (ICERs) for seven maternal retesting scenarios that included HIV testing during early antenatal care (ANC) and retesting from late ANC through nine months postpartum.

**Results:** Retesting during late ANC was estimated to avert 16% (Kenya), 14% (South Africa), and 8% (Ukraine) of infant HIV infections among key populations. Retesting during late ANC was cost-saving or cost-effective among all populations included in our model in South Africa and Kenya. In Ukraine, HIV retesting during late ANC was cost-saving for PWID but not cost-effective for FSW. Postpartum retesting was not cost-effective in any population.

**Conclusions:** Maternal HIV retesting during late ANC is cost-saving or cost-effective for vertical HIV transmission outcomes for pregnant key populations and serodiscordant couples in Kenya and South Africa and is cost-saving for PWID in Ukraine.

**Funding:** USAID, Bill and Melinda Gates Foundation, and National Institutes of Health

## INTRODUCTION

The risk of HIV transmission from pregnant and breastfeeding individuals to their children is high among those who are not on treatment and achieving viral suppression. Incident infections during pregnancy and postpartum account for nearly 30% of all vertical transmission.(1)(2) HIV retesting during pregnancy and postpartum may contribute to preventing vertical transmission when offered following initial testing in antenatal care (ANC).(3) Globally, key populations account for approximately 20% of new HIV infections and HIV incidence is 2-15 times higher among key populations than the general population, with variation by geography and key population group.(4) Pregnant and postpartum people in serodiscordant partnerships, as well as those from key populations including female sex workers (FSWs) and people who inject drugs (PWIDs), may benefit from optimized retesting strategies due to their risk of HIV acquisition. Prior model-based analyses have found HIV retesting in late pregnancy (third trimester) is cost-effective among the general population in high- and intermediate-HIV prevalence settings;(5) however, the optimal frequency and timing of HIV testing for key pregnant populations is uncertain.

Current WHO HIV testing recommendations for key populations include retesting at least annually, with more frequent testing for people with higher risk or eligible for pre-exposure prophylaxis (PrEP). For pregnant individuals, testing should take place as early as possible when ANC begins followed by retesting in the third trimester, or catch-up testing at the next possible time in settings where HIV prevalence and incidence are high.(6) To help direct limited resources toward HIV prevention and treatment goals, we assessed the health and economic impacts of different HIV retesting strategies on pregnant and breastfeeding FSW, PWID, and people in serodiscordant partnerships in three countries with differing HIV burden: South Africa (high), Kenya (intermediate), and Ukraine (low).

## METHODS

We adapted a prior Markov model designed to assess the cost-effectiveness of HIV retesting during pregnancy through 12 months postpartum among the general population.(5) We constructed separate country-specific models for FSW and PWID in Kenya, South Africa, and Ukraine and for HIV-negative pregnant people in serodiscordant partnerships in Kenya and South Africa. We modeled seven retesting scenarios: 1) late pregnancy retesting, with catch-up testing at delivery or 6 weeks postpartum if no retesting occurred during pregnancy, 2) scenario 1 + 14 weeks postpartum, 3) scenario 1 + 6 months postpartum, 4) scenario 1 + 9 months postpartum, 5) scenario 1 + 14 weeks and 6 months postpartum, 6) scenario 1 + 14 weeks and 9 months postpartum, and 7) scenario 1 + every 3 months postpartum through 9 months. In the base case scenario, we assumed HIV tests only occurred at the first ANC visit. Late pregnancy is based on each country’s recommended ANC visit schedule: gestational age of 33 weeks (Kenya), 36 weeks (South Africa), and 28 weeks (Ukraine).

We modeled infant HIV infections, infant deaths, and disability-adjusted life years (DALYs) using a one-week time step from conception through 12-months postpartum. Model parameters were obtained from published literature, expert consultations, and assumptions when data were unavailable. We included parameters specific to key populations when available and used those for the general population when not. Additional model details as well as parameters that differed from the general population in our model are included in the **Supplement Table S1**. We calculated country-specific estimates of the number of pregnant individuals in each key population by multiplying the total fertility rate by the proportion of people who represent each population. HIV prevalence and incidence were key parameters that differed between key populations and the general population in each country.

We estimated costs from the provider perspective. Incremental cost-effectiveness ratios (ICERs) were calculated by dividing the change in costs by the change in DALYs comparing each scenario to the next best alternative, and dominated strategies were eliminated from the calculations. We used the following country-specific cost-effectiveness thresholds: $500 for Kenya,(7,8) $750 for South Africa,(9) and $1,000 for Ukraine.(10) Disability adjusted life years (DALYs) and costs were estimated over a 20-year time horizon. In sensitivity analysis, we included 20% relative decreases of maternal HIV incidence and prevalence and 15% PrEP use. Additionally, we conducted one-way sensitivity analyses by increasing and decreasing seven parameters by 20% (HIV test coverage, ANC attendance, HIV prevalence, HIV incidence, ART retention at 1-year postpartum, breastfeeding avoidance among those living with HIV, and HIV test costs). This study did not meet criteria for ethics review as it only used aggregated data from previously published parameters.

## RESULTS

We found HIV retesting at late ANC (scenario 1) to be the scenario that was both the lowest cost and averted the most DALYs (**Table 1, S1-S3**). This scenario is projected to avert 16%, 14%, and 8% of total infant infections among pregnant individuals from key populations in Kenya, South Africa, and Ukraine respectively compared with the base case scenario. HIV retesting in late ANC is cost-effective for preventing vertical HIV transmission outcomes in all key populations in Kenya and South Africa, and among PWID in Ukraine, but was not cost-effective among FSW in Ukraine (**Table 1, Figure 1**).

**Table 1.** Cost-effectiveness of maternal HIV retesting scenarios in general population and key populations for Kenya, South Africa, and Ukraine. Late ANC is between 36-39 weeks of gestation. Testing offered at second ANC, or at delivery if not performed at late ANC, or at 6-week MCH visit if not performed at delivery or second ANC. ^a^ICER=incremental cost effectiveness ratio, calculated as incremental costs (in 2023 US$) divided by DALYs averted compared with the next least-costly scenario. ANC=antenatal care; PP=postpartum; DALY=disability-adjusted life-year. All general population estimates are from Meisner & Roberts et al. (2021) with costs updated to 2023 US$.

| Retesting scenario | Total infant infections | Total infections averted (%) | Total cost | Incremental cost | Total DALYs | Incremental DALYs averted | ICER<sup>a</sup> |
| --- | --- | --- | --- | --- | --- | --- | --- |
| **Kenya** |  |  |  |  |  |  |  |
| General population |  |  |  |  |  |  |  |
| No retesting | 13,484 | - | \$75,011,414 | - | 1,405,727 | - | - |
| Late ANC | 10,911 | 2,573 (19%) | \$77,977,332 | \$2,965,918 | 1,391,202 | 14,525 | \$204 |
| Late ANC + 14 weeks | 10,656 | 2,828 (21%) | \$82,624,873 | \$4,647,541 | 1,389,764 | 1,438 | \$2,623 |
| FSW |  |  |  |  |  |  |  |
| No retesting | 638 | - | \$3,140,138 | - | 19370 | - | - |
| Late ANC | 556 | 82 (13%) | \$3,084,537 | -\$55,601 | 18,908 | 463 | Cost saving |
| Late ANC + 14 weeks | 549 | 89 (14%) | \$3,134,479 | \$804 | 18,868 | 21 | \$1,268 |
| PWID |  |  |  |  |  |  |  |
| No retesting | 4 | - | \$23,992 | - | 188 | - | - |
| Late ANC | 4 | <1 (12%) | \$24,067 | \$76 | 185 | 3 | \$27 |
| Late ANC + 14 weeks | 4 | <1 (14%) | \$24,695 | \$628 | 185 | <1 | \$1,443 |
| Serodiscordant couples |  |  |  |  |  |  |  |
| No retesting | 468 | - | \$1,596,413 | - | 34,575 | - | - |
| Late ANC | 376 | 93 (20%) | \$1,645,512 | \$49,099 | 34,052 | 523 | \$94 |
| Late ANC + 14 weeks | 358 | 111 (24%) | \$1,795,569 | \$7,047 | 33,950 | 61 | \$1,473 |
| **South Africa** |  |  |  |  |  |  |  |
| General population |  |  |  |  |  |  |  |
| No retesting | 27,038 | - | \$144,227,239 | - | 906,680 | - | - |
| Late ANC | 23,838 | 3,200 (12%) | \$149,363,142 | \$6,327,227 | 888,889 | 17,792 | \$356 |
| Late ANC + 14 weeks | 23,616 | 3,421 (13%) | \$154,454,205 | \$6,271,986 | 887,657 | 1,232 | \$4,132 |
| FSW |  |  |  |  |  |  |  |
| No retesting | 401 | - | \$2,755,966 | - | 9,036 | - | - |
| Late ANC | 365 | 36 (6%) | \$2,782,571 | \$26,605 | 8,836 | 200 | \$134 |
| Late ANC + 14 weeks | 363 | 38 (6%) | \$2,821,108 | \$6,290 | 8,825 | 11 | \$3,333 |
| PWID |  |  |  |  |  |  |  |
| No retesting | 587 | - | \$3,853,344 | - | 13,094 | - | - |
| Late ANC | 545 | 46 (8%) | \$3,891,390 | \$38,046 | 12,861 | 233 | \$163 |
| Late ANC + 14 weeks | 542 | 47 (8%) | \$3,943,232 | \$8,234 | 12,847 | 9 | \$3,852 |
| Serodiscordant couples |  |  |  |  |  |  |  |
| No retesting | 641 | - | \$2,212,997 | - | 41,380 | - | - |
| Late ANC | 486 | 154 (24%) | \$2,676,441 | \$463,444 | 40,521 | 859 | \$540 |
| Late ANC + 14 weeks | 469 | 171 (27%) | \$3,119,904 | \$72,501 | 40,428 | 67 | \$4,734 |
| **Ukraine** |  |  |  |  |  |  |  |
| General population |  |  |  |  |  |  |  |
| No retesting | 71 | - | \$8,470,318 | - | 57,636 | - | - |
| Late ANC | 67 | 4 (6%) | \$9,960,198 | \$1,835,473 | 57,615 | 22 | \$87,403 |
| Late ANC + 14 weeks | 66 | 5 (7%) | \$11,345,338 | \$1,706,437 | 57,610 | 4 | \$341,287 |
| FSW |  |  |  |  |  |  |  |
| No retesting | 5 | - | \$462,767 | - | 408 | - | - |
| Late ANC | 5 | <1 (12%) | \$466,054 | \$3,287 | 405 | 3 | \$1,010 |
| Late ANC + 14 weeks | 5 | <1 (13%) | \$477,982 | \$11,928 | 404 | 0 | \$27,626 |
| PWID |  |  |  |  |  |  |  |
| No retesting | 34 | - | \$2,937,316 | - | 629 | - | - |
| Late ANC | 32 | 3 (8%) | \$2,908,757 | -\$28,559 | 615 | 14 | Cost-saving |
| Late ANC + 14 weeks | 31 | 3 (8%) | \$2,919,410 | \$40 | 615 | 0 | \$20,520 |

**Figure 1.**
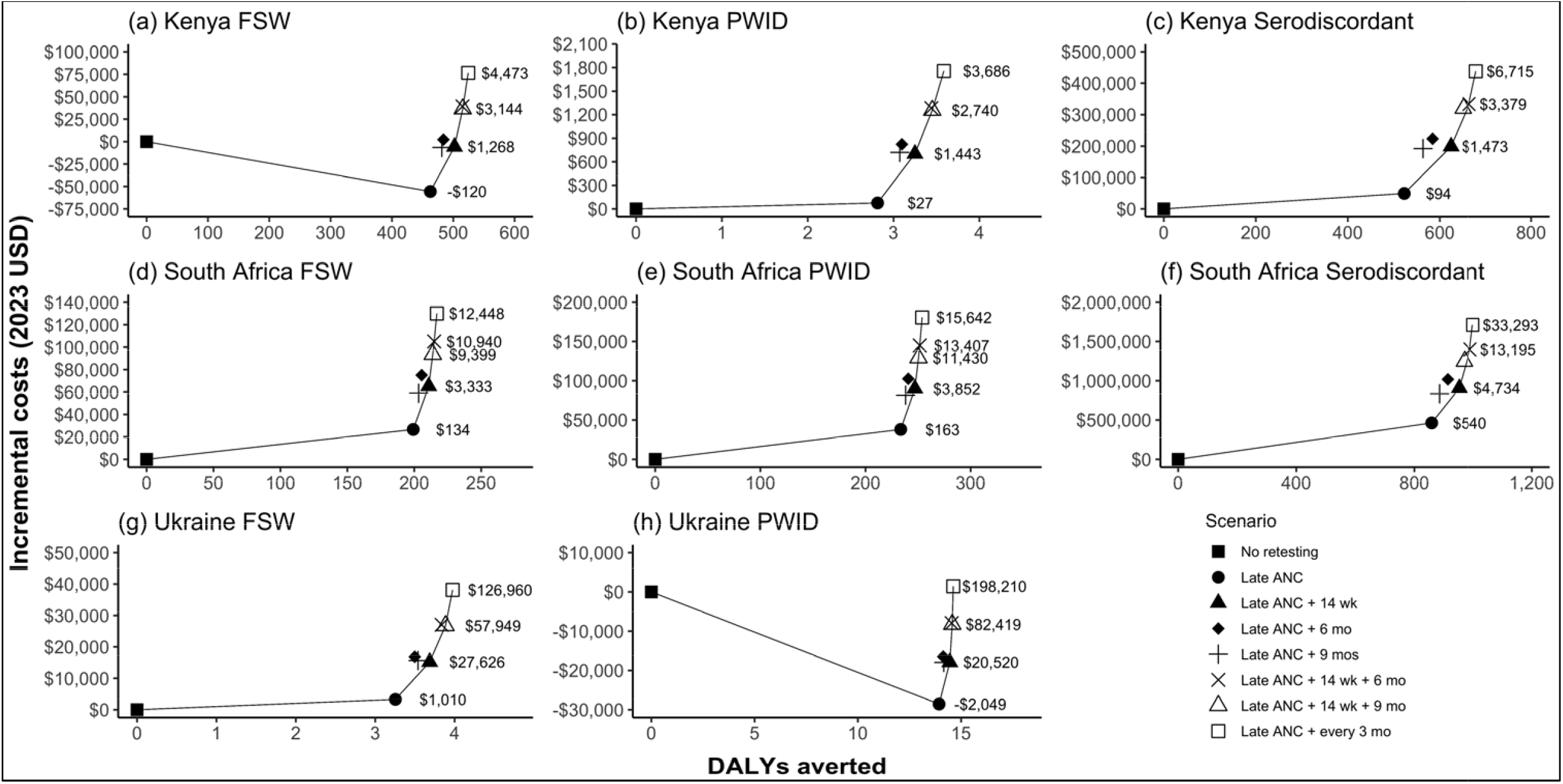
**Efficiency frontier** showing the disability adjusted life years (DALYs) averted and incremental costs (in 2023 USD) compared to the base case scenario for seven retesting scenarios. (a) Kenya FSW, (b) Kenya PWID, (c) Kenya serodiscordant couples, (d) South Africa FSW, (e) South Africa PWID, (f) South Africa serodiscordant couples, (g) Ukraine FSW, (h) Ukraine PWID. The solid line indicates scenarios that are not dominated by another, where “dominated” indicates a scenario is more costly and less effective. Note that axes are different for each graph.

Comparing impact by key population, in Kenya and South Africa, retesting in late ANC had the largest impact (by both total and percent of infections averted) among pregnant people in serodiscordant partnerships. In Ukraine, late ANC retesting averted a higher number of infections among PWID but averted a higher proportion of infections among FSW. HIV prevalence was the most important population driver for cost-effectiveness in our model among FSW in Ukraine; if HIV prevalence was 8.7% (vs. estimated 5.2%), late ANC retesting would be cost-effective in this population.

An additional retest during the postpartum period averted more DALYs but was not cost-effective for vertical HIV transmission in any scenario. Compared to retesting in late ANC alone, retesting in late ANC and once postpartum averts up to an additional 1.3%, 0.8%, and 0.5% of total infant infections in Kenya, South Africa, and Ukraine respectively, among all populations, most of which occur among people in serodiscordant partnerships. Among all populations, a postpartum test averted more infections and was more cost-effective when conducted at 14 weeks postpartum than later in the postpartum period.

Our finding that retesting in late pregnancy is cost-effective was robust to sensitivity analyses in all three countries. Decreasing HIV incidence and prevalence by 20% and adding 15% PrEP uptake increased ICERs for HIV retesting during late pregnancy for all populations while remaining below the cost-effectiveness threshold (**Supplemental Tables S4-S6**). Similarly, late ANC retesting scenarios also remained cost-effective after adjusting for seven key model parameters independently, with the exception of PWID in Ukraine where this scenario was no longer cost-effective (ICER=$1,922) when initial HIV testing coverage was increased from 97% to 100% (**Supplement Figure S3**).

## DISCUSSION

Overall, retesting in late pregnancy was cost-saving or cost-effective among key populations in Kenya and South Africa, and among PWID in Ukraine. Our finding that retesting late in pregnancy among PWID is cost-saving in Ukraine contrasts with our previous finding that HIV retesting during pregnancy among the general population was not cost-effective in Ukraine.(5) The number of model-projected HIV infections averted among PWID in Ukraine is small (3 infections per year), hence findings are subject to considerable uncertainty; however, the incremental cost of this strategy is also small ($3,287 per year). Although HIV retesting among FSW in Ukraine exceeded the cost-effective threshold, it is less costly and more effective than retesting the general population ($1,010 vs. $87,403 per DALY averted).(5) These findings suggest that HIV retesting among key populations may be cost-effective in low HIV prevalence settings where retesting pregnant people in the general population is not an efficient strategy. In Ukraine, < 60% of FSWs and PWIDs are aware of their HIV status, highlighting the need for more HIV testing among key populations in this country.(4)

Retesting every 3 months postpartum averted 20% of total infant infections in all three countries combined, suggesting additional interventions such as PrEP and partner testing, particularly among key populations, are necessary for elimination of vertical HIV transmission. Our finding that late ANC retesting was no longer cost-effective among PWID in Ukraine when early ANC HIV testing coverage reached 100% implies that these results are driven by identifying people who were previously infected, but not tested, rather than incident infections that occurred between early and late ANC, and highlights the important role of ‘catch-up’ testing in maternal retesting strategies. This also suggests that HIV retesting during late ANC for key populations in low HIV burden settings such as Ukraine may be less cost-effective with near universal early ANC testing, but still cost-effective with very high testing coverage (>99%) at early ANC.

Limited resources for HIV programs in settings with low HIV burden may make it challenging to prioritize maternal HIV retesting among key populations; however, our model found late ANC retesting to be cost-effective even with reduced ANC attendance. South African guidelines already include HIV retesting and postpartum testing every three months for all pregnant individuals,(11) but countries without additional HIV testing recommendations (or more constrained HIV testing resources) for pregnant people may need to conduct targeted outreach to improve uptake of maternal services, particularly as key populations already face added barriers to accessing reproductive health services.(12) Even in countries with guidelines for late pregnancy and postpartum testing there may be challenges to implementation; one study reports only 32% of pregnant individuals in Kenya were retested during pregnancy, but the reasons behind this are unclear.(13) Focusing retesting efforts on one specific timepoint, such as the third trimester, may be logistically simpler to implement and more accessible for all pregnant populations, compared to more complex testing algorithms based on specific risk factors.

Our study was subject to some limitations. Empirically derived parameters for people who are both pregnant and belong to high-risk populations are limited. In estimating model parameters, we assumed that pregnant people in key populations were more similar to non-pregnant people from key populations than they were to pregnant people in the general population, and we used estimates specific to key populations whenever possible. However, information on specific HIV testing, risk behavior, and population size was not available for all model parameters. Using data from the general population may underestimate the impact of maternal HIV retesting. Monitoring the changing HIV epidemiology among pregnant key populations, particularly FSW in concentrated epidemics, is important to measure progress towards elimination of vertical HIV transmission. Further, we assume pregnant people in key populations who access ANC have similar HIV risk as those without healthcare access. A limitation specific to Ukraine is that our estimates are based on pre-war populations and parameters. The ongoing Russian invasion of Ukraine that began in 2022 has impacted access to medical care through the destruction of medical facilities and displacement of healthcare workers.(14,15) This has disrupted ANC and HIV services for all people in Ukraine, including those from key populations, and represents another barrier to preventing vertical HIV transmission in the country. Finally, we do not capture benefits of preventing additional HIV infections beyond vertical HIV transmission. Treatment as prevention strategies reduce viral loads and overall transmission risk of those with HIV by facilitating linkage to ART. These benefits are likely substantial among key populations; previous work has shown that testing more frequently (every 3-6 months) can be cost-effective in concentrated epidemics in non-pregnant populations.(16,17) Models that incorporate both vertical HIV transmission and onward transmission risks among PWID and FSW may find additional postpartum tests among these populations cost-effective.

## CONCLUSIONS

HIV retesting of members of key populations during late pregnancy averted 16%, 14%, and 8% of infant infections in Kenya, South Africa, and Ukraine, respectively. This strategy is cost-effective or cost-saving among key populations in generalized epidemics (Kenya, South Africa) and may be cost-effective in concentrated epidemics (Ukraine). Retesting late in pregnancy continues to be the most cost-effective strategy, and programs with limited resources should consider prioritizing resources and efforts to optimize testing coverage, along with expanded access to early ART and PrEP, to have the greatest impact on reducing new HIV infections. Efforts are needed to focus implementation of maternal 0 retesting on those with high ongoing HIV risk.

## Data Availability

All data produced in the present work are contained in the manuscript

## Declaration of interests

The authors have no competing interests to declare. The views expressed in this manuscript are those of the authors and do not necessarily represent the official position of the WHO.

## Acknowledgement

This study was funded by WHO #201742717, WHO #018/CDS/HIV/004, WHO #2018/865307-0, USAID GHA□G□00□09□00003, and the Bill and Melinda Gates Foundation OPP1177903, and supported by NIH T32 HD10142 (DMC), NIH/NIAID P30□AI027757, NIH/NIAID K01 AI116298 (ALD), NIH/NIMH K01 MH115789 (MS), NIH/NIEHS T32 ES015459-09 (JM), and NIH/NIMH F30 MH122300 (DAR). The findings and conclusions in this report are those of the authors and do not necessarily represent the official position of the funding agencies.

## Author’s Contributions

JM, DAR, and PR developed the initial model. DC, JM, DAR, and PR parameterized the model and carried out the model implementation. MNO, MS, AB, AV, AW, GG, TC, RVB, SF, SE, MSJ, RB, CCJ, and ALD provided critical feedback and helped shape the research, analysis, and manuscript. All authors have read and approved the final manuscript.

## Notes

### Competing Interest Statement

The authors have declared no competing interest.

