## Supplemental file for "Maternal HIV retesting during pregnancy and postpartum among high-risk populations in Kenya, South Africa, and Ukraine: Cost-effectiveness of preventing vertical transmission"

**Model description**

We adapted a prior Markov model that was designed to measure the cost-effectiveness of HIV retesting during pregnancy and 12 months post-partum in Kenya, South Africa, and Ukraine. The full model details are published elsewhere.(1) Briefly, this is a deterministic state-transition model created using Excel software. There are 6 maternal HIV stages: 1 HIV negative, and 5 HIV positive states (acute HIV on ART, acute HIV not on ART, chronic HIV unknown ART status, chronic HIV on ART, chronic HIV not on ART). These 6 stages are stratified on 9 antenatal and postpartum periods – Stage 0: onset of pregnancy to first ANC; stage 1: first ANC to late ANC; stage 2: late ANC to delivery; stage 3: delivery; stage 4: first 6 weeks postpartum; stage 5: six to 14 weeks postpartum; stage 6: 14 weeks to six months postpartum; stage 7: six to nine months postpartum; stage 8: nine to 12 months postpartum. This results in a total of 54 states. Population size within each state represented in our model move between HIV negative to acute HIV positive and not on ART, and between HIV stages depending on our deterministic parameters.

Our model only includes pregnant members of key populations (FSW, PWID, HIV negative women in serodiscordant partnerships), and we estimate the number of individuals in each group using the estimated number of each key population per country and the fertility rate for each country. For our model parameters, we assume that key population parameters are the same as the general population parameters found in the original paper unless we found evidence that it differs. **Table S1** lists parameters that differ between our model and the original models. We assumed higher HIV prevalence among FSW (29%, 58%, and 5%) and among PWID (18%, 58%, and 31%) than the general population (6.1%, 31%, and 0.7%) in Kenya, South Africa, and Ukraine, respectively.(2–4) We also assumed higher HIV incidence among pregnant people in serodiscordant partnerships (7.02 per 100 PY in late pregnancy),(5) among FSW (4.7, 6.4, and 0.26 per 100 PY in Kenya, South Africa, and Ukraine respectively),(6,7) and among PWID (5.7, 6.4, and 0.35 per 100 PY in Kenya, South Africa, and Ukraine),(8,9) than the general population (1.18, 1.72, and 0.01 per 100 PY in Kenya, South Africa, and Ukraine).(5,10,11) Due to the absence of empirical HIV incidence rates for FSW in Ukraine we calculated incidence based on the country's incidence:prevalence ratio and the prevalence of HIV among FSW. We assumed HIV prevalence and incidence among PWID were the same as FSW in South Africa.

Infant disability adjusted life-years (DALYs) and treatment costs were estimated over a 20-year time horizon and discounted at 3% annually. We model the number of infant infections but estimate cost and health events based on model outputs (infant infections). The use of pre-exposure prophylaxis (PrEP) is included in a sensitivity analysis, and we incorporate it into our model through reduced incidence of HIV.

### Model parameters

**Table S1. Key model parameters.** We present only parameters that are different from the prior model. For the full list of prior model parameters see Meisner & Roberts et al. (2021). [1] Pregnant key population sizes were calculated using estimates of key population sizes and number of pregnancies per woman per country. [2] Maternal HIV incidence assumes zero risk of HIV during the period from delivery to 6 weeks postpartum. ‡ = same as non-key population for country; * = assumption. ANC=antenatal care, FSW=female sex workers, PWID=people who inject drugs, SD=Pregnant individuals in serodiscordant relationships, ART=antiretroviral therapy.

| **Parameter** | **Kenya** | | | **South Africa** | | | **Ukraine** | |
|---|---|---|---|---|---|---|---|---|
| **Key population** | **FSW** | **PWID** | **SD** | **FSW** | **PWID** | **SD** | **FSW** | **PWID** |
| Population of pregnant individuals[1] | 19,348 (12) | 201 (2) | 39,180 (13) | 9,906 (14) | 14,309 (15) | 55,035 (16) | 2,413 (17) | 2,882 (18) |
| Percent of individuals who know HIV status at onset of pregnancy | 0.68 (19) | 0.94 (2) | 0.96‡ | 0.81 (3) | 0.80 (3) | 0.94‡ (3) | 0.58 (4) | 0.43 (4) |
| *HIV risk* | | | | | | | | |
| HIV prevalence | 29.3% (2) | 18.3% (2) | 0* | 57.7% (3) | 57.7%* | 0* | 5.2% (20) | 31.4% (4) |
| Maternal HIV incidence rate from pregnancy to 12 months postpartum (per 100 person-years)[2] | 4.7 (6) | 5.70 (8) | 3.75 (5) | 6.4 (6) | 6.4* | 3.75 (5) | 0.26 (7) | 0.35 (9) |
| *Health care visits* | | | | | | | | |
| Attend first ANC | 96%‡ (17) | 96%‡ (17) | 96%‡ (17) | 90% (21) | 94%‡ (22) | 94%‡ (22) | 99%‡ (23) | 99%‡ (23) |
| First ANC (gestational age in weeks) | 22‡ (17) | 22‡ (17) | 22‡ (17) | 19 (21) | 18‡ (16) | 18‡ (16) | 10*‡ | 10*‡ |
| *Antiretroviral coverage* | | | | | | | | |
| Maternal ART initiation | 87%‡ (24) | 87%‡ (24) | 87%‡ (24) | 94% (25) | 87%‡ (24) | 87%‡ (24) | 95%‡ (7) | 88% (26) |
| Viral suppression among those on ART | 88%‡ (27) | 88%‡ (27) | 88%‡ (27) | 72%‡ (28) | 72%‡ (28) | 72%‡ (28) | 88%‡ (27) | 88%‡ (27) |
| Weekly risk of ART dropout | 0.56% (25) | 0.56% (25) | 0.56% (25) | 0.56% (25) | 0.56% (25) | 0.56% (25) | 0.56% (25) | 0.56% (25) |
| *Maternal mortality rate (per 100 person-years)* | | | | | | | | |
| During pregnancy | 0.52‡ (29) | 0.52‡ (29) | 0.52‡ (29) | 0.52‡ (29) | 0.52‡ (29) | 0.52‡ (29) | 0.10‡ (29) | 0.31 (30) |
| Delivery through 6 weeks postpartum | 4.68‡ (31) | 4.68‡ (31) | 4.68‡ (31) | 1.04‡ (31) | 1.04‡ (31) | 1.04‡ (31) | 0.21‡ (31) | 0.31 (30) |
| 6 weeks to 12 months postpartum | 0.52‡ (29) | 0.52‡ (29) | 0.52‡ (29) | 0.52‡ (29) | 0.52‡ (29) | 0.52‡ (29) | 0.10‡ (29) | 1.30 (32) |

### Cost Effectiveness Tables by Country

**Table S1. Cost-effectiveness of maternal HIV retesting scenarios for key populations in Ukraine.** Late ANC is between 36-39 weeks of gestation. Testing offered at second ANC, or at delivery if not performed at late ANC, or at 6-week MCH visit if not performed at delivery or second ANC. [a]ICER=incremental cost effectiveness ratio, calculated as incremental costs (in 2023 US$) divided by DALYs averted compared with the next least-costly scenario. ANC=antenatal care; PP=postpartum; DALY=disability-adjusted life-year. All general population estimates are from Meisner & Roberts et al. (2021). Values in parentheses are negative, indicating that the scenario is cost-saving.

| Retesting scenario | Infant infections | Total infections averted | % infections averted | Infant deaths | Total cost | Incremental cost | Total DALYs | Incremental DALYs averted | ICER |
|---|---|---|---|---|---|---|---|---|---|
| **Female sex workers (FSW)** | | | | | | | | | |
| No retesting | 5 | - | - | 21 | $462,767 | - | 408 | - | - |
| Late ANC | 5 | 1 | 12% | 21 | $466,054 | $3,287 | 405 | 3 | $1,010 |
| Late ANC + 14 wk | 5 | 1 | 13% | 21 | $477,982 | $11,928 | 404 | 0 | $27,626 |
| Late ANC + 9 mo | 5 | 1 | 13% | 21 | $478,420 | $438 | 404 | 0 | Dom |
| Late ANC + 6 mo | 5 | 1 | 12% | 21 | $479,616 | $1,196 | 404 | 0 | Dom |
| Late ANC + 14 wk + 9 mo | 5 | 1 | 14% | 21 | $489,536 | $9,920 | 404 | 0 | $57,949 |
| Late ANC + 14 wk + 6 mo | 5 | 1 | 14% | 21 | $489,809 | $273 | 404 | 0 | Dom |
| Late ANC + every 3 mo | 5 | 1 | 14% | 21 | $500,893 | $11,084 | 404 | 0 | $126,960 |
| **People who inject drugs (PWID)** | | | | | | | | | |
| No retesting | 34 | - | - | 29 | $2,937,316 | - | 629 | - | - |
| Late ANC | 32 | 3 | 8% | 29 | $2,908,757 | -$28,559 | 615 | 14 | ($2,049) |
| Late ANC + 9 mo | 31 | 3 | 8% | 29 | $2,919,370 | $10,612 | 615 | 0 | Dom |
| Late ANC + 14 wk | 31 | 3 | 8% | 29 | $2,919,410 | $40 | 615 | 0 | $20,520 |
| Late ANC + 6 mo | 32 | 3 | 8% | 29 | $2,920,838 | $1,428 | 615 | 0 | Dom |
| Late ANC + 14 wk + 9 mo | 31 | 3 | 8% | 29 | $2,929,074 | $8,236 | 615 | 0 | $82,419 |
| Late ANC + 14 wk + 6 mo | 31 | 3 | 8% | 29 | $2,929,376 | $302 | 615 | 0 | Dom |
| Late ANC + every 3 mo | 31 | 3 | 8% | 29 | $2,938,744 | $9,368 | 615 | 0 | $198,210 |

**Table S2. Cost-effectiveness of maternal HIV retesting scenarios for key populations in Kenya.** Late ANC is between 36-39 weeks of gestation. Testing offered at second ANC, or at delivery if not performed at late ANC, or at 6-week MCH visit if not performed at delivery or second ANC. [a]ICER=incremental cost effectiveness ratio, calculated as incremental costs (in 2017 US$) divided by DALYs averted compared with the next least-costly scenario. ANC=antenatal care; PP=postpartum; DALY=disability-adjusted life-year. All general population estimates are from Meisner & Roberts et al. (2021). Values in parentheses are negative, indicating that the scenario is cost-saving.

| Retesting scenario | Infant infections | Total infections averted | % infections averted | Infant deaths | Total cost | Incremental cost | Total DALYs | Incremental DALYs averted | ICER |
|---|---|---|---|---|---|---|---|---|---|
| **Female sex workers (FSW)** | | | | | | | | | |
| No retesting | 638 | - | - | 810 | $3,140,138 | - | 19,370 | - | - |
| Late ANC | 556 | 82 | 13% | 797 | $3,084,537 | -$55,601 | 18,908 | 463 | ($120) |
| Late ANC +9mo | 553 | 85 | 13% | 796 | $3,133,675 | $49,138 | 18,889 | 19 | Dom |
| Late ANC +14wk | 549 | 89 | 14% | 796 | $3,134,479 | $804 | 18,868 | 21 | $1,268 |
| Late ANC +6mo | 552 | 86 | 13% | 796 | $3,142,423 | $7,944 | 18,887 | -18 | Dom |
| Late ANC +14wk/9mo | 547 | 91 | 14% | 795 | $3,176,636 | $34,213 | 18,855 | 31 | $3,144 |
| Late ANC +14wk/6mo | 547 | 91 | 14% | 795 | $3,179,443 | $2,807 | 18,855 | 0 | Dom |
| Late ANC +every 3mo | 545 | 93 | 15% | 795 | $3,216,649 | $37,206 | 18,846 | 9 | $4,473 |
| **People who inject drugs (PWID)** | | | | | | | | | |
| No retesting | 4 | - | - | 8 | $23,992 | - | 188 | - | - |
| Late ANC | 4 | 0 | 12% | 8 | $24,067 | $76 | 185 | 3 | $27 |
| Late ANC +14wk | 4 | 1 | 14% | 8 | $24,695 | $628 | 185 | 0 | $1,443 |
| Late ANC +9mo | 4 | 1 | 13% | 8 | $24,712 | $17 | 185 | 0 | Dom |
| Late ANC +6mo | 4 | 1 | 13% | 8 | $24,815 | $103 | 185 | 0 | Dom |
| Late ANC +14wk/9mo | 4 | 1 | 14% | 8 | $25,248 | $433 | 184 | 0 | $2,740 |
| Late ANC +14wk/6mo | 4 | 1 | 14% | 8 | $25,275 | $28 | 184 | 0 | Dom |
| Late ANC +every 3mo | 4 | 1 | 15% | 8 | $25,748 | $472 | 184 | 0 | $3,686 |
| **Serodiscordant couples** | | | | | | | | | |
| No retesting | 468 | - | - | 1,504 | $1,596,413 | - | 34,575 | - | - |
| Late ANC | 376 | 93 | 20% | 1,489 | $1,645,512 | $49,099 | 34,052 | 523 | $94 |
| Late ANC +9mo | 369 | 100 | 21% | 1,488 | $1,788,522 | $143,010 | 34,011 | 41 | Dom |
| Late ANC +14wk | 358 | 111 | 24% | 1,486 | $1,795,569 | $7,047 | 33,950 | 61 | $1,473 |
| Late ANC +6mo | 365 | 104 | 22% | 1,487 | $1,819,843 | $24,274 | 33,990 | -40 | Dom |
| Late ANC +14wk/9mo | 353 | 115 | 25% | 1,485 | $1,916,918 | $97,074 | 33,923 | 68 | Dom |
| Late ANC +14wk/6mo | 351 | 118 | 25% | 1,485 | $1,930,447 | $13,530 | 33,910 | 13 | $3,379 |
| Late ANC +every 3mo | 348 | 120 | 26% | 1,485 | $2,034,520 | $104,073 | 33,895 | 15 | $6,715 |

**Table S3. Cost-effectiveness of maternal HIV retesting scenarios for key populations in South Africa.** Late ANC is between 36-39 weeks of gestation. Testing offered at second ANC, or at delivery if not performed at late ANC, or at 6-week MCH visit if not performed at delivery or second ANC. [a]ICER=incremental cost effectiveness ratio, calculated as incremental costs (in 2017 US$) divided by DALYs averted compared with the next least-costly scenario. ANC=antenatal care; PP=postpartum; DALY=disability-adjusted life-year. All general population estimates are from Meisner & Roberts et al. (2021). Values in parentheses are negative, indicating that the scenario is cost-saving.

| Retesting scenario | Infant infections | Total infections averted | % infections averted | Infant deaths | Total cost | Incremental cost | Total DALYs | Incremental DALYs averted | ICER |
|---|---|---|---|---|---|---|---|---|---|
| **Female sex workers (FSW)** | | | | | | | | | |
| No retesting | 401 | | - | 395 | $2,755,966 | - | 9,036 | - | - |
| Late ANC | 365 | 36 | 6% | 389 | $2,782,571 | $26,605 | 8,836 | 199 | $134 |
| Late ANC + 9mo | 364 | 37 | 6% | 389 | $2,814,819 | $32,248 | 8,832 | 4 | Dom |
| Late ANC + 14wk | 363 | 37 | 6% | 389 | $2,821,108 | $6,290 | 8,825 | 8 | $3,333 |
| Late ANC + 6mo | 364 | 38 | 6% | 389 | $2,831,010 | $9,902 | 8,830 | -5 | Dom |
| Late ANC + 14 wk + 9mo | 362 | 39 | 7% | 389 | $2,849,540 | $18,530 | 8,822 | 8 | $9,399 |
| Late ANC + 14 wk + 6mo | 362 | 38 | 7% | 389 | $2,860,970 | $11,430 | 8,821 | 1 | $10,940 |
| Late ANC + every 3 mo | 362 | 39 | 7% | 389 | $2,885,935 | $24,966 | 8,819 | 2 | $12,448 |
| **People who inject drugs (PWID)** | | | | | | | | | |
| No retesting | 587 | - | - | 572 | $3,853,344 | - | 13,094 | - | - |
| Late ANC | 545 | 46 | 8% | 565 | $3,891,390 | $38,046 | 12,861 | 233 | $163 |
| Late ANC + 9mo | 544 | 47 | 8% | 565 | $3,934,998 | $43,609 | 12,856 | 5 | Dom |
| Late ANC + 14wk | 542 | 47 | 8% | 565 | $3,943,232 | $8,234 | 12,847 | 9 | $3,852 |
| Late ANC + 6mo | 543 | 49 | 8% | 565 | $3,955,899 | $12,667 | 12,854 | -6 | Dom |
| Late ANC + 14 wk + 9mo | 542 | 50 | 8% | 564 | $3,982,636 | $26,737 | 12,844 | 10 | $11,430 |
| Late ANC + 14 wk + 6mo | 541 | 50 | 8% | 564 | $3,998,350 | $15,713 | 12,843 | 1 | $13,407 |
| Late ANC + every 3 mo | 541 | 50 | 9% | 564 | $4,033,953 | $35,603 | 12,840 | 2 | $15,642 |
| **Serodiscordant couples** | | | | | | | | | |
| No retesting | 641 | - | - | 1940 | $2,212,997 | - | 41,380 | - | - |
| Late ANC | 486 | 154 | 24% | 1,915 | $2,676,441 | $463,444 | 40,521 | 859 | $540 |
| Late ANC + 9mo | 482 | 159 | 25% | 1,914 | $3,047,403 | $370,962 | 40,495 | 27 | Dom |
| Late ANC + 14wk | 469 | 171 | 27% | 1,913 | $3,119,904 | $72,501 | 40,428 | 67 | $4,734 |
| Late ANC + 6mo | 476 | 164 | 26% | 1,914 | $3,231,467 | $111,563 | 40,467 | -39 | Dom |
| Late ANC + 14 wk + 9mo | 466 | 175 | 27% | 1,912 | $3,462,528 | $231,061 | 40,409 | 57 | Dom |
| Late ANC + 14 wk + 6mo | 463 | 178 | 28% | 1,911 | $3,611,918 | $149,390 | 40,391 | 19 | $13,195 |
| Late ANC + every 3 mo | 461 | 180 | 28% | 1,911 | $3,924,983 | $313,065 | 40,381 | 9 | $33,293 |

**Sensitivity analyses**

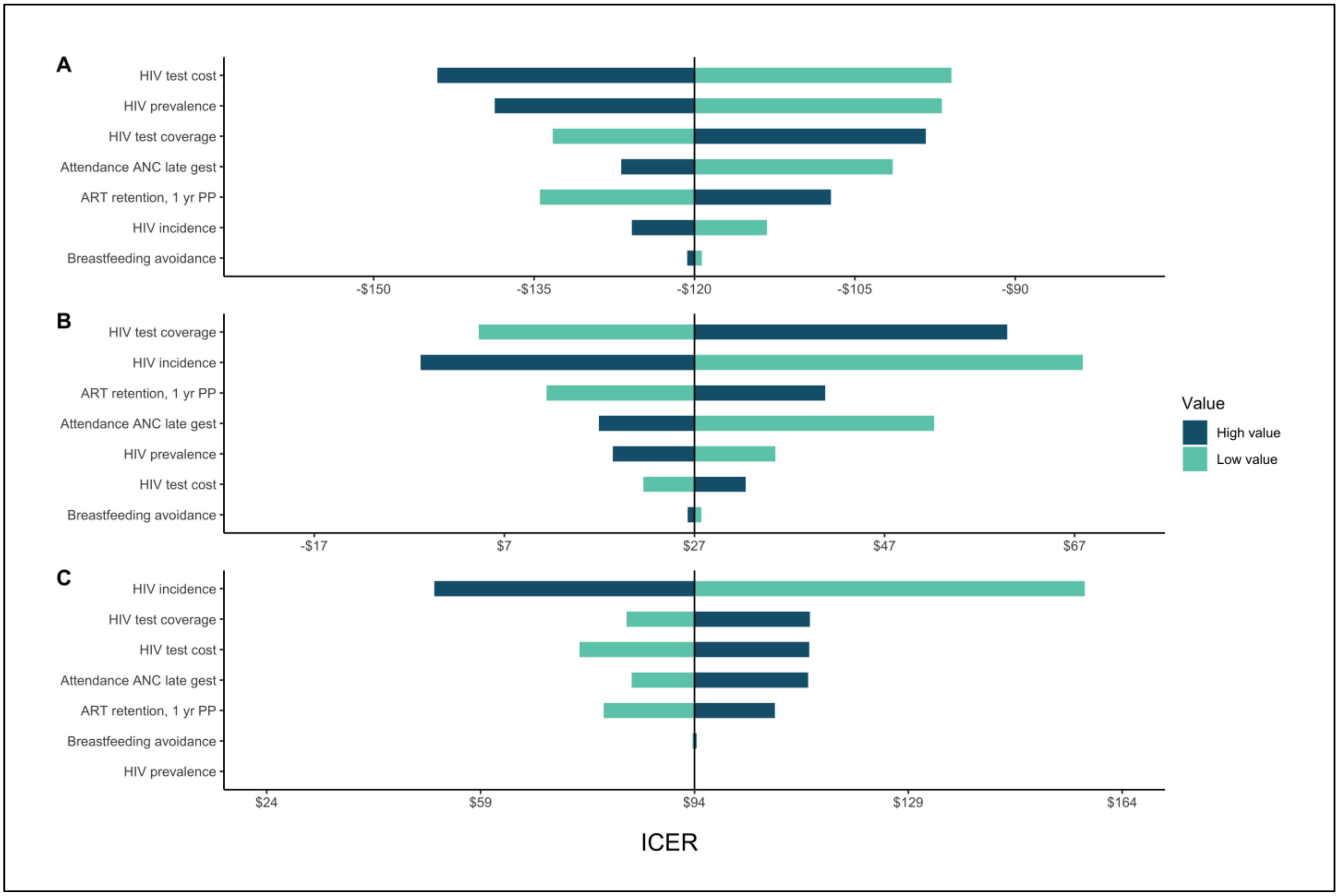

**Figure S1. One-way sensitivity analysis for Kenya late ANC retesting: (A) FSW, (B) PWID, (C) Serodiscordant couples.** Each model parameter (y-axis) was individually adjusted 20% higher (High value) and 20% lower (low value). The x-axis shows the change in ICER from the unadjusted value. Each population is centered on the unadjusted ICER. Note that x-axes are at different scales. ANC = Antenatal care

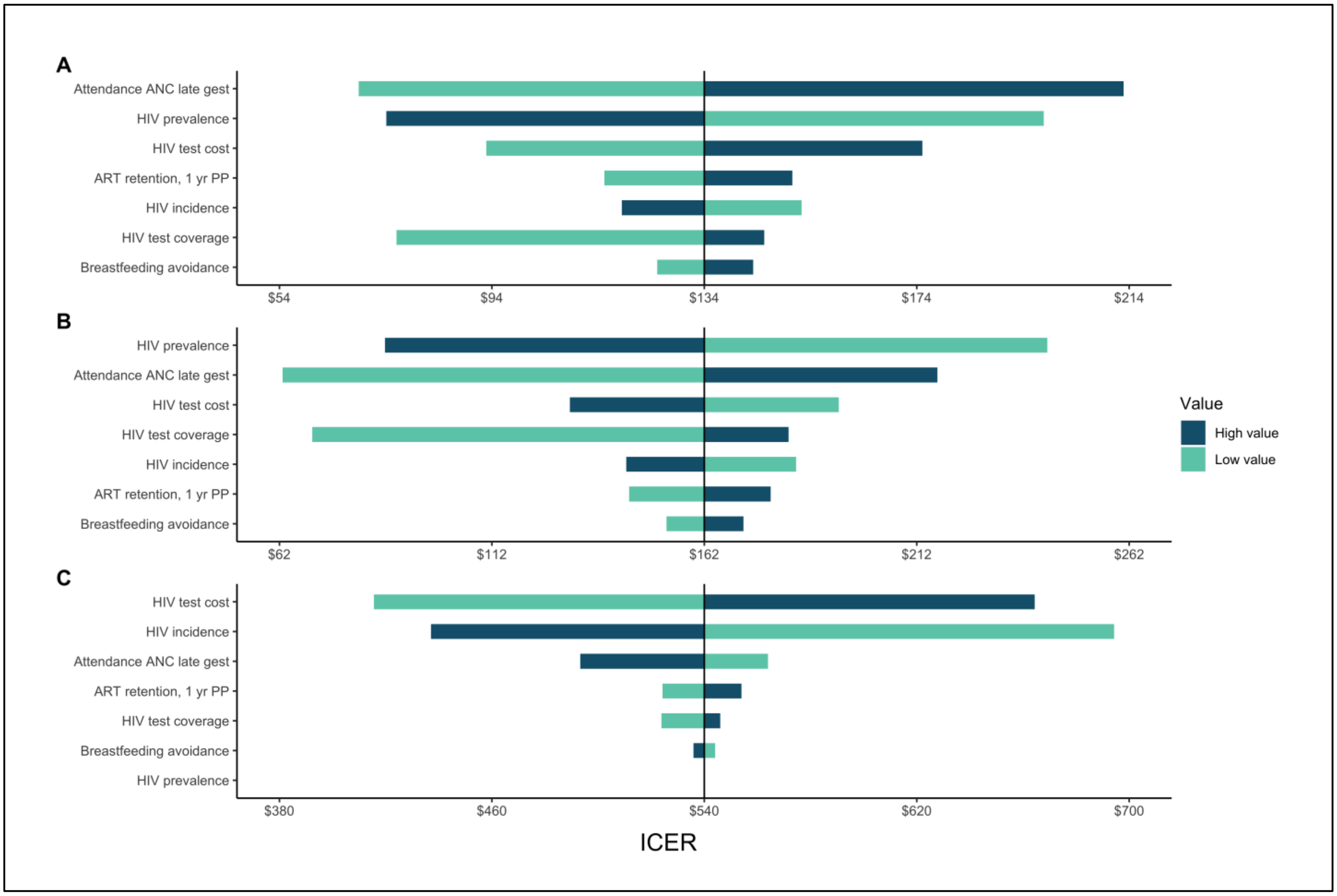

**Figure S2. One-way sensitivity analysis for South Africa late ANC retesting: (A) FSW, (B) PWID, (C) Serodiscordant couples.** Each model parameter (y-axis) was individually adjusted 20% higher (High value) and 20% lower (low value). The x-axis shows the change in ICER from the unadjusted value. Each population is centered on the unadjusted ICER. Note that x-axes are at different scales. ANC = Antenatal care

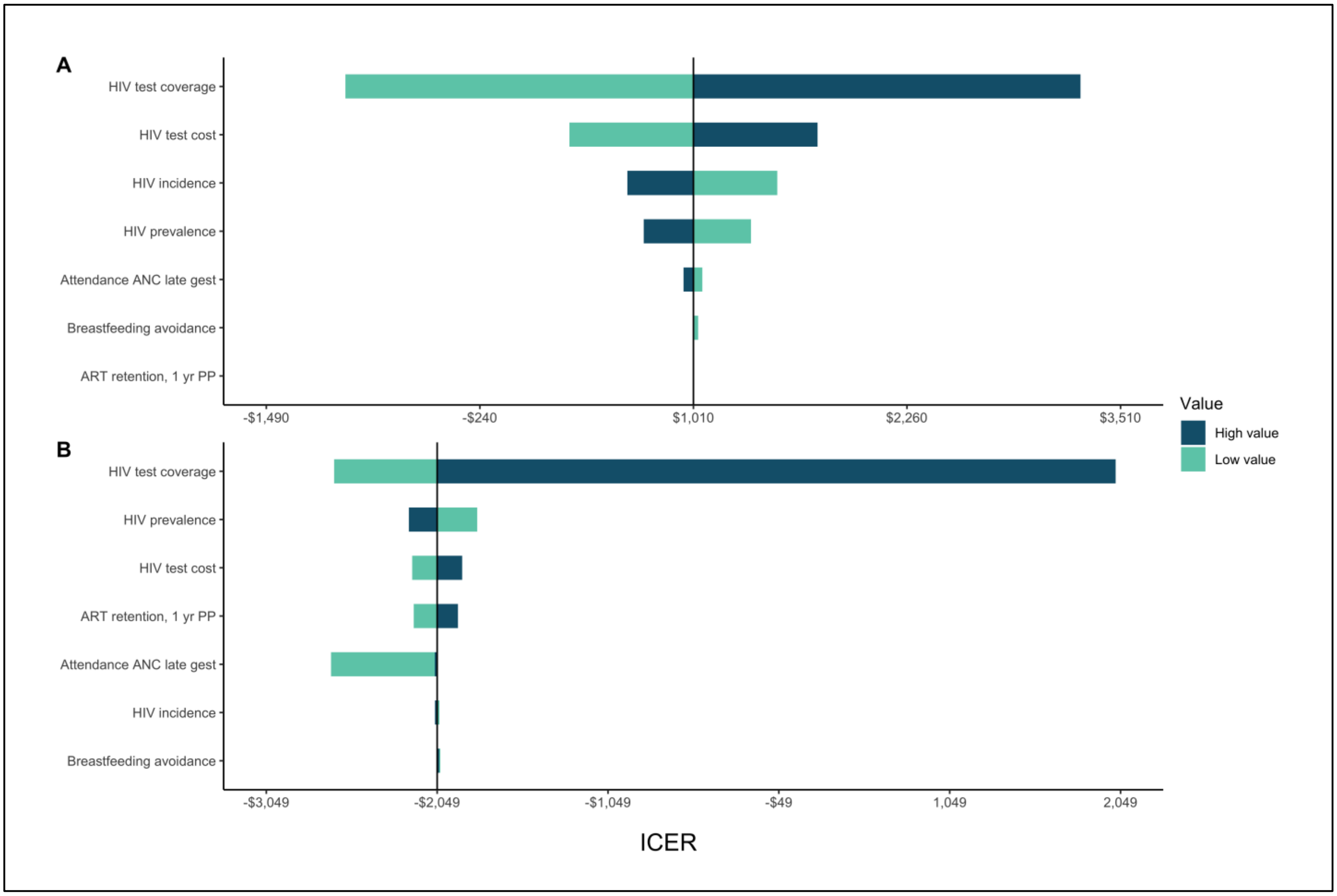

**Figure S3. One-way sensitivity analysis for Ukraine late ANC retesting: (A) FSW, (B) PWID.** Each model parameter (y-axis) was individually adjusted 20% higher (High value) and 20% lower (low value). The x-axis shows the change in ICER from the unadjusted value. Each population is centered on the unadjusted ICER. Note that x-axes are at different scales. ANC = Antenatal care

**Table S4. Cost-effectiveness of maternal HIV retesting scenarios for key populations in Ukraine: sensitivity analysis including 20% decreases in HIV incidence and prevalence, and 15% PrEP uptake.** Late ANC is between 36-39 weeks of gestation. Testing offered at second ANC, or at delivery if not performed at late ANC, or at 6-week MCH visit if not performed at delivery or second ANC. [a]ICER=incremental cost effectiveness ratio, calculated as incremental costs (in 2023 US$) divided by DALYs averted compared with the next least-costly scenario. ANC=antenatal care; PP=postpartum; DALY=disability-adjusted life-year. All general population estimates are from Meisner & Roberts et al. (2021). Values in parentheses are negative, indicating that the scenario is cost-saving.

| Retesting scenario | Infant infections | Total infections averted | % infections averted | Infant deaths | Total cost | Incremental cost | Total DALYs | Incremental DALYs averted | ICER |
|---|---|---|---|---|---|---|---|---|---|
| **Female sex workers (FSW)** | | | | | | | | | |
| No retesting | 5 | - | - | 21 | $1,210,247 | - | 407 | - | - |
| Late ANC | 5 | 1 | 12% | 21 | $1,214,013 | $3,766 | 404 | 3 | $1,229 |
| Late ANC + 14 wk | 5 | 1 | 13% | 21 | $1,225,811 | $11,798 | 404 | 0 | $30,313 |
| Late ANC + 9 mo | 5 | 1 | 13% | 21 | $1,226,220 | $409 | 404 | 0 | Dom |
| Late ANC + 6 mo | 5 | 1 | 13% | 21 | $1,227,310 | $1,090 | 404 | 0 | Dom |
| Late ANC + 14 wk + 9 mo | 5 | 1 | 14% | 21 | $1,237,285 | $9,976 | 404 | 0 | $64,310 |
| Late ANC + 14 wk + 6 mo | 5 | 1 | 14% | 21 | $1,237,543 | $258 | 404 | 0 | Dom |
| Late ANC + every 3 mo | 5 | 1 | 14% | 21 | $1,248,598 | $11,055 | 403 | 0 | $141,361 |
| **People who inject drugs (PWID)** | | | | | | | | | |
| No retesting | 34 | - | - | 29 | $3,578,237 | - | 629 | - | - |
| Late ANC | 31 | 3 | 8% | 29 | $3,550,023 | -$28,214 | 615 | 14 | ($2,044) |
| Late ANC + 9 mo | 31 | 3 | 8% | 29 | $3,560,543 | $10,520 | 615 | 0 | Dom |
| Late ANC + 14 wk | 31 | 3 | 8% | 29 | $3,560,595 | $52 | 614 | 0 | $21,588 |
| Late ANC + 6 mo | 31 | 3 | 8% | 29 | $3,561,939 | $1,344 | 615 | 0 | Dom |
| Late ANC + 14 wk + 9 mo | 31 | 3 | 8% | 29 | $3,570,217 | $8,279 | 614 | 0 | $91,214 |
| Late ANC + 14 wk + 6 mo | 31 | 3 | 8% | 29 | $3,570,508 | $290 | 614 | 0 | Dom |
| Late ANC + every 3 mo | 31 | 3 | 8% | 29 | $3,579,860 | $9,353 | 614 | 0 | $220,855 |

**Table S5. Cost-effectiveness of maternal HIV retesting scenarios for key populations in Kenya: sensitivity analysis including 20% decreases in HIV incidence and prevalence, and 15% PrEP uptake.** Late ANC is between 36-39 weeks of gestation. Testing offered at second ANC, or at delivery if not performed at late ANC, or at 6-week MCH visit if not performed at delivery or second ANC. [a]ICER=incremental cost effectiveness ratio, calculated as incremental costs (in 2017 US$) divided by DALYs averted compared with the next least-costly scenario. ANC=antenatal care; PP=postpartum; DALY=disability-adjusted life-year. All general population estimates are from Meisner & Roberts et al. (2021). Values in parentheses are negative, indicating that the scenario is cost-saving.

| Retesting scenario | Infant infections | Total infections averted | % infections averted | Infant deaths | Total cost | Incremental cost | Total DALYs | Incremental DALYs averted | ICER |
|---|---|---|---|---|---|---|---|---|---|
| **Female sex workers (FSW)** | | | | | | | | | |
| No retesting | 593 | - | - | 803 | $4,385,208 | - | 19,115 | - | - |
| Late ANC | 518 | 74 | 13% | 790 | $4,338,338 | -$46,871 | 18,695 | 420 | ($112) |
| Late ANC +9mo | 516 | 77 | 13% | 790 | $4,383,789 | $45,452 | 18,681 | 14 | Dom |
| Late ANC +14wk | 513 | 80 | 13% | 789 | $4,385,283 | $1,493 | 18,663 | 17 | $1,476 |
| Late ANC +6mo | 516 | 77 | 13% | 790 | $4,391,055 | $5,773 | 18,678 | -15 | Dom |
| Late ANC +14wk/9mo | 511 | 82 | 14% | 789 | $4,425,058 | $34,003 | 18,653 | 25 | $3,991 |
| Late ANC +14wk/6mo | 511 | 82 | 14% | 789 | $4,427,486 | $2,428 | 18,653 | 0 | Dom |
| Late ANC +every 3mo | 510 | 83 | 14% | 789 | $4,463,489 | $36,003 | 18,646 | 7 | $5,755 |
| **People who inject drugs (PWID)** | | | | | | | | | |
| No retesting | 4 | - | - | 8 | $39,789 | - | 187 | - | - |
| Late ANC | 4 | 0 | 11% | 8 | $39,894 | $106 | 184 | 3 | $41 |
| Late ANC +14wk | 4 | 1 | 13% | 8 | $40,497 | $603 | 184 | 0 | $1,533 |
| Late ANC +9mo | 4 | 1 | 12% | 8 | $40,510 | $13 | 184 | 0 | Dom |
| Late ANC +6mo | 4 | 1 | 12% | 8 | $40,604 | $94 | 184 | 0 | Dom |
| Late ANC +14wk/9mo | 3 | 1 | 14% | 8 | $41,031 | $427 | 184 | 0 | $2,934 |
| Late ANC +14wk/6mo | 3 | 1 | 14% | 8 | $41,056 | $26 | 184 | 0 | Dom |
| Late ANC +every 3mo | 3 | 1 | 14% | 8 | $41,517 | $461 | 183 | 0 | $3,983 |
| **Serodiscordant couples** | | | | | | | | | |
| No retesting | 425 | - | - | 1,497 | $5,429,103 | - | 34,332 | - | - |
| Late ANC | 341 | 84 | 20% | 1,483 | $5,484,704 | $55,601 | 33,858 | 473 | $117 |
| Late ANC +9mo | 335 | 90 | 21% | 1,482 | $5,622,472 | $137,768 | 33,822 | 37 | Dom |
| Late ANC +14wk | 325 | 100 | 24% | 1,481 | $5,628,709 | $6,237 | 33,767 | 55 | $1,572 |
| Late ANC +6mo | 332 | 94 | 22% | 1,482 | $5,650,266 | $21,557 | 33,803 | -36 | Dom |
| Late ANC +14wk/9mo | 321 | 104 | 25% | 1,480 | $5,747,007 | $96,740 | 33,742 | 61 | Dom |
| Late ANC +14wk/6mo | 319 | 106 | 25% | 1,480 | $5,758,859 | $11,852 | 33,731 | 11 | $3,627 |
| Late ANC +every 3mo | 316 | 109 | 26% | 1,479 | $5,861,683 | $102,824 | 33,717 | 14 | $7,376 |

**Table S6. Cost-effectiveness of maternal HIV retesting scenarios for key populations in South Africa: sensitivity analysis including 20% decreases in HIV incidence and prevalence, and 15% PrEP uptake.** Late ANC is between 36-39 weeks of gestation. Testing offered at second ANC, or at delivery if not performed at late ANC, or at 6-week MCH visit if not performed at delivery or second ANC. [a]ICER=incremental cost effectiveness ratio, calculated as incremental costs (in 2017 US$) divided by DALYs averted compared with the next least-costly scenario. ANC=antenatal care; PP=postpartum; DALY=disability-adjusted life-year. All general population estimates are from Meisner & Roberts et al. (2021). Values in parentheses are negative, indicating that the scenario is cost-saving.

| Retesting scenario | Infant infections | Total infections averted | % infections averted | Infant deaths | Total cost | Incremental cost | Total DALYs | Incremental DALYs averted | ICER |
|---|---|---|---|---|---|---|---|---|---|
| **Female sex workers (FSW)** | | | | | | | | | |
| No retesting | 383 | - | - | 392 | $3,120,617 | - | 8,937 | - | - |
| Late ANC | 351 | 36 | 6% | 387 | $3,148,859 | $28,242 | 8,758 | 179 | $158 |
| Late ANC + 9mo | 350 | 38 | 7% | 387 | $3,179,280 | $30,421 | 8,755 | 3 | Dom |
| Late ANC + 14wk | 349 | 37 | 6% | 387 | $3,185,629 | $6,349 | 8,749 | 6 | $3,903 |
| Late ANC + 6mo | 350 | 37 | 6% | 387 | $3,194,241 | $8,612 | 8,753 | -5 | Dom |
| Late ANC + 14 wk + 9mo | 349 | 38 | 7% | 387 | $3,212,909 | $18,668 | 8,747 | 7 | $12,036 |
| Late ANC + 14 wk + 6mo | 349 | 39 | 7% | 387 | $3,223,949 | $11,040 | 8,746 | 1 | $13,623 |
| Late ANC + every 3 mo | 348 | 39 | 7% | 387 | $3,248,560 | $24,612 | 8,744 | 1 | $16,586 |
| **People who inject drugs (PWID)** | | | | | | | | | |
| No retesting | 581 | - | - | 571 | $4,439,821 | - | 13,061 | - | - |
| Late ANC | 540 | 46 | 8% | 564 | $4,478,396 | $38,575 | 12,835 | 226 | $171 |
| Late ANC + 9mo | 539 | 47 | 8% | 564 | $4,521,151 | $42,754 | 12,831 | 4 | Dom |
| Late ANC + 14wk | 538 | 47 | 8% | 564 | $4,529,425 | $8,274 | 12,822 | 8 | $4,062 |
| Late ANC + 6mo | 539 | 49 | 8% | 564 | $4,541,519 | $12,094 | 12,828 | -6 | Dom |
| Late ANC + 14 wk + 9mo | 537 | 50 | 9% | 564 | $4,568,242 | $26,723 | 12,819 | 9 | $12,425 |
| Late ANC + 14 wk + 6mo | 537 | 50 | 9% | 564 | $4,583,756 | $15,514 | 12,818 | 1 | $14,419 |
| Late ANC + every 3 mo | 537 | 50 | 9% | 564 | $4,619,101 | $35,345 | 12,816 | 2 | $17,214 |
| **Serodiscordant couples** | | | | | | | | | |
| No retesting | 574 | | - | 1929 | $7,607,327 | | 41,007 | | - |
| Late ANC | 434 | 139 | 24% | 1,907 | $8,076,000 | $468,673 | 40,233 | 774 | $606 |
| Late ANC + 9mo | 431 | 143 | 25% | 1,906 | $8,434,616 | $358,616 | 40,212 | 22 | Dom |
| Late ANC + 14wk | 420 | 153 | 27% | 1,905 | $8,507,021 | $72,405 | 40,154 | 58 | $5,432 |
| Late ANC + 6mo | 427 | 147 | 26% | 1,906 | $8,606,414 | $99,393 | 40,190 | -36 | Dom |
| Late ANC + 14 wk + 9mo | 418 | 156 | 27% | 1,904 | $8,841,382 | $234,969 | 40,139 | 51 | Dom |
| Late ANC + 14 wk + 6mo | 415 | 159 | 28% | 1,904 | $8,983,480 | $142,098 | 40,126 | 14 | $16,834 |
| Late ANC + every 3 mo | 414 | 160 | 28% | 1,904 | $9,294,850 | $311,370 | 40,118 | 8 | $41,068 |
